# Coronavirus in pregnancy and delivery: rapid review and expert consensus

**DOI:** 10.1101/2020.03.06.20032144

**Authors:** E Mullins, D Evans, RM Viner, P O’Brien, E Morris

**Affiliations:** Imperial College; RCPCH; RCOG

**Keywords:** Pregnancy, Pregnant, Coronavirus, SARS, Severe acute respiratory syndrome, MERS Middle East Respiratory Syndrome, HCov-229E, HCov-NL63, HCov-OC43, HCov-HKU1

## Abstract

**BACKGROUND:** Person to person spread of COIVD-19 in the UK has now been confirmed. There are limited case series reporting the impact on women affected by coronaviruses (CoV) during pregnancy. In women affected by SARS and MERS, the case fatality rate appeared higher in women affected in pregnancy compared with non-pregnant women. We conducted a rapid, review to guide management of women affected by COVID -19 during pregnancy and developed interim practice guidance with the RCOG and RCPCH to inform maternity and neonatal service planning

**METHODS:** Searches were conducted in PubMed and MedRxiv to identify primary case reports, case series, observational studies or randomised-controlled trial describing women affected by coronavirus in pregnancy and on neonates. Data was extracted from relevant papers and the review was drafted with representatives of the RCPCH and RCOG who also provided expert consensus on areas where data were lacking

**RESULTS:** From 9964 results on PubMed and 600 on MedRxiv, 18 relevant studies (case reports and case series) were identified. There was inconsistent reporting of maternal, perinatal and neonatal outcomes across case reports and series concerning COVID-19, SARS, MERS and other coronaviruses. From reports of 19 women to date affected by COVID-19 in pregnancy, delivering 20 babies, 3 (16%) were asymptomatic, 1 (5%) was admitted to ICU and no maternal deaths have been reported. Deliveries were 17 by caesarean section, 2 by vaginal delivery, 8 (42%) delivered pre-term. There was one neonatal death, in 15 babies who were tested there was no evidence of vertical transmission.

**CONCLUSIONS:** Morbidity and mortality from COVID-19 appears less marked than for SARS and MERS, acknowledging the limited number of cases reported to date. Pre-term delivery affected 42% of women hospitalised with COVID-19, which may put considerable pressure on neonatal services if the UK reasonable worse-case scenario of 80% of the population affected is realised. There has been no evidence of vertical transmission to date. The RCOG and RCPCH have provided interim guidance to help maternity and neonatal services plan their response to COVID-19.

## BACKGROUND

The common human coronaviruses; 1. 229E (alpha coronavirus), 2. NL63 (alpha coronavirus), 3. OC43 (beta coronavirus) and 4. HKU1 (beta coronavirus) cause the common cold. Three human coronaviruses cause more severe, acute illnesses; MERS-CoV causes Middle East Respiratory Syndrome, MERS, SARS-CoV causes severe acute respiratory syndrome, or SARS and <u>SARS-CoV-2 coronavirus that causes COVID-1</u>9.

There are limited case series reporting the impact on women affected by coronaviruses (CoV) during pregnancy. In women affected by SARS and MERS, the case fatality rate appeared higher in women affected in pregnancy compared with non-pregnant women.

Person to person spread of COIVD-19 in the UK has now been confirmed. To guide treatment and prevention in women affected COVID-19 during pregnancy in the current outbreak we conducted a rapid review to guide management, of women affected by COVID -19 during pregnancy. Expert consensus will be required in areas where data is lacking at this early stage in the COVID-19 outbreak.

## METHODS

Searches were conducted in PubMed and MedRxiv, see appendix for search strategies, to identify primary case reports, case series or randomised-controlled trial describing women of any age affected by coronavirus in pregnancy or the postnatal period. There were no date or language restrictions to the search. References of relevant papers were hand searched for relevant studies.

Due to time constraints one person, EM, conducted the search, reviewed full texts and extracted data on demographics, maternal outcomes, maternal diagnostic testing, maternal imaging, fetal outcomes, perinatal outcomes, neonatal outcomes and neonatal diagnostic testing. Subjective comparison of outcomes for COVID-19, SARS and MERS is presented.

The initial draft of review was drafted with representatives of the RCPCH and RCOG who also provided expert consensus on areas where data were lacking. This is presented as an appendix to this review. Registration on PROSPERO and contact with study authors was not undertaken due to time constraints. Quality of study was assessed subjectively as anecdotal, low, medium or high.

## RESULTS

***PubMed search*** There were 9964 search results, 68 abstracts were screened, 50 were excluded as not including pregnant women or humans or being in-vitro studies. Eighteen relevant studies were identified, their full texts were reviewed and all 18 were included. It is highly likely that there were overlaps in cases reported to be affected by SARS. ***MedRxiv search*** 600 results, 39 abstracts were screened, no relevant studies identified.

There was inconsistent reporting of maternal, perinatal and neonatal outcomes across case reports and series. All studies were case reports or series with varying levels of detail on the data and outcomes assessed, subjectively all were of low quality. A narrative review is presented.

**Table 1.** Overview of pregnancy, perinatal and neonatal outcomes in pregnancies affected by coronaviruses

|  | COVID-19 | SARS |  |  | MERS |  |  |
| --- | --- | --- | --- | --- | --- | --- | --- |
| **N** | 19 (20 infants) | 20 |  |  | 11 |  |  |
| Median age (range) | 30 (25-40) |  |  |  |  |  |  |
| Median gestational age at presentation (range) | 36.5 (31-39) | 16 (3-32) |  |  | 24 (6-38) |  |  |
| Symptomatic at admission | 16 (84%) | 20 (100%) |  |  | 9 (81%) |  |  |
| ICU admission (%) | 1* | 6 (30%) |  |  | 7 (63%) |  |  |
| Maternal death (%) | 0* | 3 (15%) |  |  | 3 (27%) |  |  |
| CT-chest/CXR changes in symptomatic women | 17 (94%) | 20 (100%) |  |  | 8/9*** (89%) |  |  |
| Stage of pregnancy | 3rd Trimester | 1st trimester | 2nd Trimester | 3rd Trimester | 1st Trimester | 2nd Trimester | 3rd Trimester |
| **N** | 19 (20 infants) | 7 | 5 | 8 (9 fetuses) | 1 | 5 | 5 |
| Women with co-morbidities | 4 (21%) | not reported | not reported | not reported | 0 | 2 (40%) | 3 (60%) |
| Admitted asymptomatic | 3 (16%) | 0 | 0 | 0 | 1 (100%) | 1 (20%) | 0 |
| ICU admission % | 1 (5%) | 1 (14%) | 2 (40%) | 3 (38%) | 0 | 3 (60%) | 4 (80%) |
| Maternal mortality % | 0* | 1 (14%) | 1 (20%) | 1 (13%) | 0 | 1 (20%) | 2 (40%) |
| Miscarriage or intra-uterine death | 0 | 4 (58%) | 1 (20%) | 1 (1 twin) (13%) | 0 | 1 (20%) | 1 (20%) |
| Any pre-term delivery | 8/19 (42%)* | not reported | 2 (40%) | 2 (26%) | 0 | 1 (20%) | 2 (40%) |
| Spontaneous pre-term delivery | 0 | not reported | 0** | 1 (13%)* | 0 | 0 | 0 |
| Iatrogenic pre-term delivery | 3/10 (30%)* | not reported | 2 (40%)* | 1 (13%)* | 0 | 1 (20%) | 2 (40%) |
| Fetal growth restriction post-infection | 0 | not reported | 0 | 2 (26%) | 0 | 0 | 0 |
| Vertical transmission | 0 | 0 | 0 | 0 | 0 | 0 | 0 |
| Neonatal death | 1/20 (5%) | not reported | 0 | 0 | 0 | 1 (20%) | 0 |

\* Incomplete data from Zhu et al
\*\* Incomplete data from Zhang et al. n=5
\*\*\*One woman declined radiography because of concerns about effect on pregnancy
ICU-Intensive Care Unit, CT-computed tomography, CXR - chest X-ray

## MATERNAL OUTCOMES

### COVID 19

So far, 19 women affected in pregnancy have been reported, delivering 20 infants (1–3). 17 were delivered by caesarean and 2 by vaginal delivery, all within 13 days of onset of illness.Where reported maternal morbidity and mortality (n=10), 1 woman required ICU and mechanical ventilation, all women had viral changes apparent on CT chest imaging and there were no maternal deaths (1,2).

### SARS

Case fatality rate (CFR) was 15% for all reported cases of SARS in pregnancy (4–9). A case-control study comparing 10 pregnant and 40 non-pregnant women affected by SARS in Hong Kong reported 60% ICU admission and 40% CFR in the pregnant group compared with 17.5% and 0% in the non-pregnant group (7). All women affected by SARS had CT or chest X-ray evidence of pneumonia.

### MERS

In pregnant women affected by MERS, 7/11 (63%) were admitted to ICU and CFR was 3/11 (27%) (10–15).

**Table 2.**
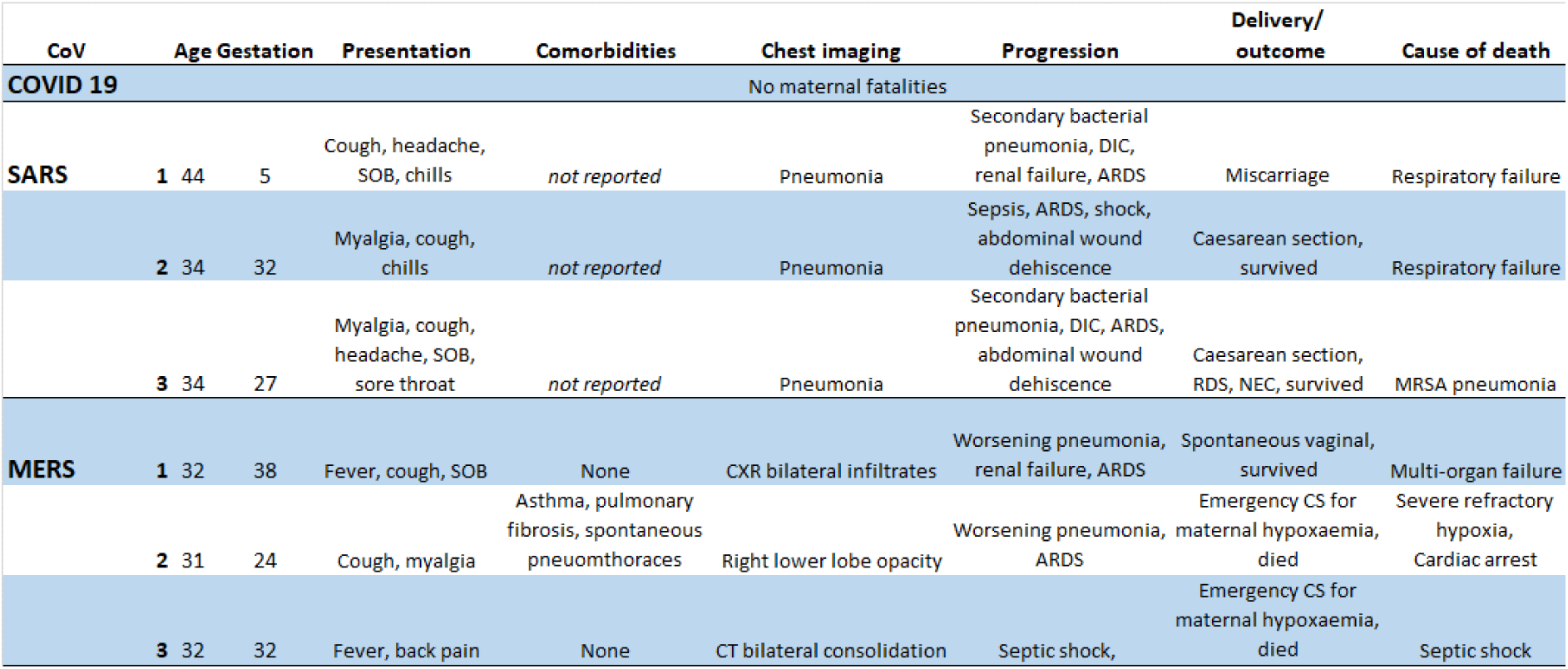
Details of women affected by coronavirus in pregnancy who died

## EARLY PREGNANCY

### COVID 19

No data to date.

### SARS

Miscarriage affected 4/7 women with 1^st^ trimester SARS infection (6). These 4 women all had ultrasound findings of 3-5 week pregnancies of unknown location or unknown viability in which ongoing pregnancy at 13 weeks would be expected in 38% and 50% respectively (acknowledging the complexity in this area (16,17)). Those with fetal heart activity recorded (n=2) did not miscarry, neither did a woman in whom the diagnosis was retrospective and did not undergo ultrasound scan.

### MERS

a single case of a women with MERS has been reported in the first trimester. This woman was asymptomatic and went on to have a term delivery (18).

## PREGNANCY LOSS

### COVID-19

There have been no pregnancy losses in the cases reported to date.

### SARS

In cases reported after the 1^st^ trimester, Zhang et al reported a series of 5 women affect (2 in 2^nd^ trimester, 3 in 3^rd^ trimester) the loss of one twin with the other surviving to delivery. It is not clear if this was in the 2^nd^ or 3^rd^ trimester, this has been nominally recorded as being in the third trimester(19).

### MERS

Three pregnancy losses were reported:

1. Became ill at 19 weeks gestation and experienced vaginal bleeding resulting in a late fetal loss at 20 weeks(15). It should be noted that this woman refused chest radiography and medications because of her concerns about their effect on pregnancy.
2. Presented at 34 weeks with pre-eclampsia and MERS and was found to have had an intra-uterine death; this woman delivered vaginally and recovered after ICU admission without ventilatory support(12).
3. A woman with a background history of pulmonary fibrosis, asthma and spontaneous pneumothoraces presented at 24 weeks gestation with MERS pneumonia requiring ICU and ventilator support. She developed ARDS and underwent emergency caesarean section to improve maternal oxygenation, the infant weighed 240g and died four hours after birth, the woman died 4 weeks after this with severe refractory hypoxia and cardiac arrest(12).

## PREMATURITY

### COVID 19

8/19 (42%) women affected delivered pre-term. In Chen et al, n=9, all mothers were electively delivered by caesarean section, 2 at 36 weeks gestation. In Zhu et al, 7 delivered by Caesarean sections and 2 by vaginal delivery, 5/9 women (6/10 babies) delivered pre-term. The indication for delivery is not reported however 6 babies were affected by fetal distress prior to delivery-it seems reasonable to assume that fetal condition contributed. Wang et al report one woman who was delivered at 30 weeks for fetal distress(2).

### SARS

4/16 pregnancies not affected by miscarriage resulted in pre-term delivery, at 26, 28, 32 and 33 weeks gestation(18), data on timing of delivery is not reported in the series of 5 women from Zhang et al(19).

### MERS

3/9 pregnancies not affected by stillbirth or intrauterine death were delivered pre-term by caesareans section, 1 at 24 weeks and 2 at 32 weeks for maternal hypoxaemia.

## FETAL GROWTH AND PLACENTAL EFFECTS

### COVID-19

Delivery occurred in all women within 13 days of onset of illness, fetal growth is unlikely to be affected in this time period. No placental pathology is available to date.

### SARS

show early changes also seen in pregnancies with fetal growth restriction (fibrin deposition) when delivery ≤1 week after onset illness, birthweight was normal in these pregnancies. When delivery was 5-7 weeks after illness onset there was fetal growth restriction in 2/3 ongoing pregnancies and their placentas showed more severe changes (areas with loss of blood supply, avascular villi, and bleeding behind placenta, abruption).

### MERS

4/11 women went on to deliver healthy babies at term, although birth weights are not reported for 3/4 of these. In one, a vaginal bleeding was reported at 37 weeks, causing fetal compromise and necessitating emergency caesarean section resulting in the delivery of a male infant, weighing 3140g in good condition. Abruption was apparent on placental examination (11).

## DELIVERY AND POSTNATAL

### COVID 19

Chen et al. reports 9 women delivering by caesarean section from 36 weeks onwards, 2 pre-term. In two women at term, fetal distress was reported. In 6 of these women with COVID-19 who delivered by caesarean section and underwent testing after delivery by caesarean section, there was no evidence of SARS-CoV 2 in amniotic fluid, cord blood neonatal throat swab or breastmilk samples(1). A news report of a baby with a COVID-19 infected mother testing positive at 30 hours has not been reported in scientific journals.

Zhu et al report COVID 19 in 9 women delivering 10 infants, 7 by caesarean section and 2 by vaginal delivery, 3 of which only became symptomatic after delivery. The indication for delivery was not reported. This cohort had COVID-19 from 31 weeks onwards, 6/9 pregnancies showed fetal distress and 5/9 women (6/10 babies) were delivered prematurely (3).

Wang et al reported on one woman who underwent caesarean section for fetal distress at 30 weeks gestation, the infant was born in good condition and samples of amniotic fluid, neonatal gastric samples, placenta and infant throat swabs were negative for SARS-CoV 2 (2).

No vertical transmission was reported for cases of ***SARS*** or ***MERS*** in pregnancy after caesarean or vaginal delivery.

### Other Coronaviruses

A single case series reported 3 out of 7 neonates born to mothers positive for HCoV 229E had HCoV 229E on RT-PCR in gastric samples; seroconversion was not assessed, no signs of infant infection were seen in those with positive gastric samples (20)

## NEONATAL OUTCOMES

### COVID-19

In Chen et al, n=9, all babies were delivered after 36 weeks and were well at discharge (1). Zhu et al reported a cohort born at earlier gestation (from 31 weeks gestation), 6/10 babies were admitted to the NNU for respiratory support, 2 developed DIC and 1 multiple organ failure (3). Neonatal morbidity was more marked in this series probably due to greater prematurity, one baby died after being born at 34 weeks. Requiring admission at 30 minutes of life with respiratory difficulties, the baby deteriorated, developed shock, DIC and multi organ failure and dies at 8 days of life. 9/10 infants were tested for COVID-19, all testing was negative. Wang et al reported a baby born at 30 weeks in good condition with an uneventful neonatal course(2).

### SARS

4/16 pregnancies not affected by miscarriage resulted in pre-term delivery, at 26, 28, 32 and 33 weeks gestation (4,6,7,19,21). The baby born at 26 weeks had respiratory distress syndrome (RDS) and a bowel perforation. The baby born at 28 weeks had RDS, necrotising enterocolitis and a patent ductus arteriosus(6,9).

### MERS

3/9 pregnancies not affected by stillbirth or intrauterine death were delivered pre-term by caesareans section, 1 at 24 weeks resulted in a neonatal death (birthweight 240g), 2 babies delivered at 32 weeks for maternal hypoxaemia have no outcomes reported (10,18).

## CONCLUSIONS

There are limited data on the impact of the current COVID-19 outbreak on women affected in pregnancy and their babies. Studies were all case-reports or series and of low quality. Outcomes reported varied, one series on COVID-19 did not report on maternal outcomes.

From the 10/19 women where maternal outcomes were reported, severe morbidity and mortality are not evident in mothers as they were for SARS and MERS.

From the data, an increase in risk of miscarriage cannot be ruled out at this stage given the SARS data. Data from early pregnancy units are needed on women affected by COVID-19 with matched controls.

In women affected by COVID-19 with ongoing pregnancy, for surveillance for fetal growth restriction would be reasonable, given the acute and chronic placental changes observed and 2/3 ongoing pregnancies being affected by fetal growth restriction after SARS and an abruption noted after MERS.

The need for provision for fetal monitoring including serial ultrasound for women with COVID-19 will be challenging for maternity services. Women will need to be cared for in units with appropriate neonatal intensive care facilities as COVID-19 is associated with pre-term delivery in 50% of reported cases.

In SARS and MERS-affected cases, delivery was most often indicated by maternal hypoxaemia. In COVID-19, if maternal illness is not as severe, the considerations will be based more on obstetric indications for delivery.

Information on vertical transmission for COVID-19 is limited, although testing of 15 neonates born to mothers with COVID-19 has all been negative. Guidance on mode of delivery requires expert consensus until further information emerges.

There is evidence for vertical transmission of HCoV 229E, seroconversion was not investigated however and all infants remained well, but there is no evidence for vertical transmission for any other coronavirus.

Consensus guidance from China advises “delayed cord clamping is not recommended” in order to reduce the risk of vertical transmission and that infants should be separated from mothers affected by COVID 19(22). Infants may acquire COVID-19 from their mothers after delivery via normal routes of transmission.

Guidance from China is that “Infants should not be fed with the breast milk from mothers with confirmed or suspected of 2019-nCoV.” Guidance from the CDC is less clear but is still precautionary (23).

Expert consensus is needed for breastfeeding and the use of expressed breastmilk for infant feeding for mothers affected by COVID-19 until more informative data emerges and for deciding whether women with COVID-19 who are symptomatic, suggesting high viral load, should breastfeed babies with unknown status.

If the UK reasonable worse-case scenario of 80% attack rate for the population is realised and 4% require hospitalisation, thousands of women during pregnancy will be affected at a time when staff will themselves be unwell. In previous coronavirus epidemics there has anecdotally been a tendency towards admitting any symptomatic, pregnant woman with proven infection.

A surge in workload will likely be seen in the NHS and across the world at a time when staffing is well below optimal levels. Pragmatic choices will need to be made about achievable and acceptable levels of care with national guidance and local adaptation. It will need to be emphasised that chest imaging should be undertaken as clinically indicated in pregnant women.

Therapeutics announced as being under consideration and trial in the outbreak include Kaletra (Lopinavir and Ritonavir), Remdesivir and Chloroquine. Kaletra (24) is used in the UK during pregnancy for treatment of HIV where the benefits of treatment outweigh the risks of toxicity seen in animal studies. The use of Chloroquine outweighs the risks in the prevention and treatment of malaria during pregnancy (25). Remdesivir has been used for the treatment of Ebola in pregnant women(26), however it should be acknowledged that Ebola is a condition with a CFR of 50% for which there would be higher tolerance for adverse effects of a potentially beneficial treatment than would be the case for COVID-19 where the CFR is around 1%. It would seem reasonable not to exclude seriously ill pregnant women from trials of these therapies for COVID-19.

There is a need for systematic data reporting on women affected by COVID-19 and their pregnancies to provide an evidence base for management, treatment and prevention and to target limited resources during the outbreak.

## APPENDICES

### I) RCOG and RCPCH joint advice for care of women affected by COVID 19 in pregnancy, full practice guidance is in draft at present and as data emerges will be updated

#### 1. Antenatal care

- *We concur that while numbers remain small, all cases of mothers known to have COVID-19 be referred to and managed in specialist centres with appropriate expertise in fetal medicine and co-located neonatal intensive care. As numbers of women affected by COVID 19 in pregnancy increase, it is likely to become necessary to care for women locally with referral where appropriate for fetal growth restriction, pre-term delivery and women requiring treatment on an Infectious Diseases specialist unit or HCID unit*
- *In common with other maternal infections, decisions about the use of corticosteroids for fetal lung maturity should be made in consultation with infectious disease, neonatal and maternal-fetal medicine consultants*

#### 2. Delivery

- *Any guidance must be made in the absence of almost any useful information. Therefore the precautionary principle should guide us, but must balance risks to mother and baby*.
- *Decision on the mode of delivery should be individualised. Caesarean section may be necessitated by maternal hypoxaemia or fetal growth restriction*.
- *We note the very recent consensus guidance from China suggests that “delayed cord clamping (DCC) is not recommended” in order to reduce the risk of vertical transmission. We recommend that the baby can be cleaned of maternal blood and secretions quickly and delayed cord clamping can be undertaken*.

#### 3. Care of mother and newborn

- *Separation of infected mother and newborn is recommended by China and US-CDC advice is that this should be considered. We do not concur that mothers and baby should be routinely separated. We believe that routine precautionary separation of mother and baby is not in the best interests of either party, in the current state of knowledge. This is particularly true given the prolonged infectious period. We note those babies most at risk, i*.*e. very preterm or those with respiratory compromise, will be on NICU and therefore necessarily separated from mothers. We emphasise that this might need to change as knowledge grows*.
- *Infants born to those with COVID-19 need to be closely monitored. The RCPCH British Paediatric Surveillance Unit (BPSU) presents opportunities for surveillance*.

#### 4. Infant feeding

- *Given the lack of data on vertical transmission, guidance on breastfeeding is difficult. We note that guidance from China is that “Infants should not be fed with the breast milk from mothers with confirmed or suspected of 2019-nCoV*.*” Guidance from the CDC is less clear but suggests being precautionary*.
- *In the current state of knowledge, we recommend that mothers with proven infection and who are symptomatic (suggestive of high viral load) should not breastfeed babies with unknown status currently. If these mothers wish to breastfeed in the future, they should express milk and discard it and resume breastfeeding once confirmed as non-infective*.
- *In less severe cases we recommend that whether to start or continue breastfeeding should be determined by the mother, in coordination with her family and healthcare providers*.
- *We recommend that human breast milk banks should not use human milk from mothers confirmed to be infected with COVID-19*

### II) Search Strategies

#### PubMed

(“gravidity”[MeSH Terms] OR “gravidity”[All Fields] OR “pregnant”[All Fields]) OR (“pregnancy”[MeSH Terms] OR “pregnancy”[All Fields]) OR (“breast feeding”[MeSH Terms] OR “breast feeding”[All Fields] OR “breastfeeding”[All Fields]) OR lactating[All Fields] OR (“infant, newborn”[MeSH Terms] OR (“infant”[All Fields] AND “newborn”[All Fields]) OR “newborn infant”[All Fields] OR “neonatal”[All Fields]) AND SARS[All Fields] OR (“severe acute respiratory syndrome”[MeSH Terms] OR “severe acute respiratory syndrome”[All Fields]) OR (“coronavirus infections”[MeSH Terms] OR (“coronavirus”[All Fields] AND “infections”[All Fields]) OR “coronavirus infections”[All Fields] OR “mers”[All Fields]) OR ((“middle east”[MeSH Terms] OR (“middle”[All Fields] AND “east”[All Fields]) OR “middle east”[All Fields]) AND acute[All Fields] AND respiratory[All Fields] AND (“syndrome”[MeSH Terms] OR “syndrome”[All Fields])) OR (coronaviral[All Fields] OR coronavirdae[All Fields] OR coronaviren[All Fields] OR coronavirida[All Fields] OR (“coronaviridae”[MeSH Terms] OR “coronaviridae”[All Fields]) OR coronaviridea[All Fields] OR (“coronaviridae”[MeSH Terms] OR “coronaviridae”[All Fields] OR “coronavirinae”[All Fields]) OR coronavirion[All Fields] OR coronavirions[All Fields] OR (“coronavirus”[MeSH Terms] OR “coronavirus”[All Fields]) OR coronavirus’[All Fields] OR coronavirus’s[All Fields] OR coronavirusapplikation[All Fields] OR coronaviruscpe[All Fields] OR coronaviruse[All Fields] OR (“coronavirus”[MeSH Terms] OR “coronavirus”[All Fields] OR “coronaviruses”[All Fields]) OR coronaviruses’[All Fields] OR coronavirusinfektion[All Fields] OR coronavirusinfektionen[All Fields] OR coronaviruslike[All Fields] OR coronavirusreplikation[All Fields] OR coronavirussen[All Fields] OR coronavirusurile[All Fields]) OR COVID-19[All Fields] AND “humans”[MeSH Terms]

#### MedRxiv

## Data Availability

This was a review. We are happy to share data, please email EM on

## Declarations

### Contributions

EM conceived the article, conducted the literature review and wrote the first draft. RV, DE, EM and PO’B wrote the first draft of the RCOG and RCPCH interim guidance. All authors contributed to the final draft and guidance. EM is the guarantor of this article.

### Transparency declaration

EM affirms that the manuscript is an honest, accurate, and transparent account of the study being reported; that no important aspects of the study have been omitted; and that any discrepancies from the study as planned (and, if relevant, registered) have been explained.

### Copyright

The Corresponding Author has the right to grant on behalf of all authors and does grant on behalf of all authors, an exclusive licence (or non exclusive for government employees) on a worldwide basis to the BMJ Publishing Group Ltd to permit this article (if accepted) to be published in BMJ editions and any other BMJPGL products and sublicences such use and exploit all subsidiary rights, as set out in our licence.

### Ethical approval

was not required for this review

### Competing interest statements

– EM has applied for a UKRI/MRC grant to study COVID-19 in pregnancy. No other authors have COIs to declare.

### Funding

EM’s salary was from the NIHR. There was no specific funding for this study.

### Patient and Public involvement

was not undertaken for this review

### Data sharing

We are happy to share data, please email EM on

## Notes

### Competing Interest Statement

EM has applied for a UKRI grant to study COVID-19 in pregnancy

## REFERENCES

1. Chen H, Guo J, Wang C, Luo F, Yu X, Zhang W, et al. Clinical characteristics and intrauterine vertical transmission potential of COVID-19 infection in nine pregnant women?: a retrospective review of medical records. 2020;6736(20):1–7.

2. Wang X, Zhou Z, Jianping Z, Zhu F, Tang Y, Shen X. A case of 2019 Novel Coronavirus in a pregnant woman with preterm delivery. Clin Infect Dis. 2020;

3. Zhu H, Wang L, Fang C, Peng S, Zhang L, Chang G, et al. Clinical analysis of 10 neonates born to mothers with 2019-nCoV pneumonia. 2020;9(1):51–60.

4. Robertson CA, Lowther SA, Birch T, Tan C, Sorhage F, Stockman L, et al. SARS and Pregnancy?: A Case Report. 2004;10(2):345–8.

5. Li AM, Ng PC. Severe acute respiratory syndrome (SARS) in neonates and children. 2005;461– 5.

6. Wong SF, Chow KM, Leung TN, Ng WF, Ng TK, Shek CC, et al. Pregnancy and perinatal outcomes of women with severe acute respiratory syndrome. 200292–7.

7. Lam CM, Wong F, Leung N, Chow M. A case-controlled study comparing clinical course and outcomes of pregnant and non-pregnant women with severe acute respiratory syndrome. 2004;111(August):771–4.

8. Swartz D, Graham A. Potential Maternal and Infant Outcomes from Coronavirus 2019-nCoV (SARS-CoV-2) Infecting Pregnant Women: Lessons from SARS, MERS, and Other Human Coronavirus Infections. Viruses. 2020;1–16.

9. Shek CC, Ng PC, Fung GPG, Cheng FWT, Chan PKS, Peiris MJS, et al. Infants Born to Mothers With Severe Acute Respiratory Syndrome. 2020;112(4).

10. Alserehi H, Wali G, Alshukairi A, Alraddadi B. Impact of Middle East Respiratory Syndrome coronavirus (MERS ‐CoV) on pregnancy and perinatal outcome. BMC Infect Dis [Internet]. 2016;1–4. Available from: http://dx.doi.org/10.1186/s12879-016-1437-y

11. Jeong SY, Sung SI, Sung J, Ahn SY, Kang E, Chang YS, et al. MERS-CoV Infection in a Pregnant Woman in Korea. 2017;3:5–8.

12. Assiri A, Abedi G, Malak M, Abdulaziz B, Gerber S, Watson JT. Pregnancy?: A Report of 5 Cases From Saudi Arabia. Clin Infect Dis. 2016;63(7):951–3.

13. Park MH, Kim HR, Choi DH, Sung JH, Kim JH. Emergency cesarean section in an epidemic of the middle east respiratory syndrome. 2016;1–5.

14. Malik A, Medhat K, Masry E, Ravi M, Sayed F. Middle East Respiratory Syndrome Coronavirus during Pregnancy, Abu Dhabi, United Arab Emirates, 2013. 2016;22(3):515–7.

15. Payne D, Ibrahim I, Sultan A. Stillbirth During Infection With Middle East Respiratory Syndrome Coronavirus. J Infect Dis. 2014;209(12):1870–2.

16. Bottomley C. A model and scoring system to predict outcome of intrauterine pregnancies of uncertain viability.

17. Bignardi T, Condous G, Kirk E, Van Calsters B, Van Huffel S, Timmerman D, et al. Viability of intrauterine pregnancy in women with pregnancy of unknown location?: prediction using human chorionic gonadotropin ratio vs. progesterone. Ultrasound Obs Gynecol. 2010;35:656–61.

18. Alfaraj S, Al-Twfiq J, Memish Z. Middle East Respiratory Syndrome Coronavirus (MERS-CoV) infection during pregnancy?: Report of two cases & review of the literature. J Microbiol. 2019;52:501–3.

19. Zhang J, Wang Y, Chen L, Zhang R, Xie Y. Clinical analysis of pregnancy in second and third trimesters complicated severe acute respiratory syndrome. Zhonghua Fu Chan Ke Za Zhi. 2003;38:516–20.

20. Gagneur A, Dirson E, Audebert S, Vallet S. Materno-fetal transmission of human coronaviruses?: a prospective pilot study. 2008;(August 2005):863–6.

21. Yudin M, Steele D, Sgro M, Read S, Kopplin P, Gough K. Severe acute respiratory syndrome in pregnancy. Obs Gynecol. 2005;105:124–7.

22. Wang L, Shi Y, Xiao T, Fu J, Feng X, Mu D, et al. Chinese expert consensus on the perinatal and neonatal management for the prevention and control of the 2019 novel coronavirus infection (First edition). 2020;8(3):1–8.

23. CDC. Interim Considerations for Infection Prevention and Control of Coronavirus Disease 2019 (COVID-19) in Inpatient Obstetric Healthcare Settings [Internet]. [cited 2020 Feb 28]. Available from: https://www.cdc.gov/coronavirus/2019-ncov/hcp/inpatient-obstetric-healthcare-guidance.html

24. BNF: Kaletra [Internet]. Available from: https://bnf.nice.org.uk/drug/lopinavir-with-ritonavir.html

25. BNF: CHloroquine {Internet}. Available from: https://bnf.nice.org.uk/drug/chloroquine.html#indicationsAndDoses

26. Mulangu S, Dodd L, Davey R, Mbaya O, Proschan M, Mukadi D, et al. A Randomized, Controlled Trial of Ebola Virus Disease Therapeutics. NEJM. 2019;381(24):2293–2203.

